# International impact of large multi-centre surgical trials of arthroscopic subacromial decompression

**DOI:** 10.1101/2021.11.10.21266128

**Authors:** James A. Smith, Kristin Kostka, David J. Beard, Andrew J. Carr, Jonathan L. Rees, Daniel Prieto-Alhambra

## Abstract

**Objectives:** To examine temporal trends in incidence of arthroscopic subacromial decompression (ASAD) surgery internationally during conduct and after publication of placebo controlled trials finding no evidence of meaningful benefit of ASAD for shoulder impingement.

**Design:** Observational study of incidence rates.

**Setting:** Large routinely collected datasets were used: outpatient data from Belgium and UK, and insurance claims and outpatient data from US. UK data were from Clinical Practice Research Datalink and Belgium and US data were from IQVIA. US and UK data spanned 2005 – 2019 and Belgium data 2011 – 2019.

**Participants:** Patients were eligible for inclusion in the study if they had at least one visit recorded in the database in a given year and cases were defined as patients undergoing ASAD for the first time in their records in a given year.

**Outcome measures:** We calculated incidence of ASAD over time, overall and stratified by age and sex. Characteristics of patients undergoing ASAD were also assessed over time.

**Results:** UK incidence has fallen since a peak of 4.7 per 10,000 person years in 2011 (when the CSAW trial began) to 1.8 in 2019. US incidence shows no clear pattern and remains consistently higher than the UK, at 11.5 per 100,000 person years in 2019. Changes in incidence patterns were similar across different age groups and sexes. The number of cases in Belgium was too small for meaningful conclusions.

**Conclusions:** We found ASAD rates have fallen in the UK during conduct and after publication of two large surgical RCTs from the UK and Finland that questioned the effectiveness of ASAD for shoulder impingement. A similar impact on clinical practice has not been seen in US. Further work to understand the barriers or concerns preventing international uptake of high quality evidence into clinical practice is needed.

**Strengths and limitations of this study:**

- This is the most comprehensive study of ASAD incidence we are aware of. Routinely collected datasets were used to assess proportions of the patients undergoing this procedure in several countries
- Standardised case definitions were used across databases to increase comparability of findings
- Temporal changes in database coverage and quality of reporting can influence findings. The observed variation in ASAD incidence may not be entirely attributable to changes in ASAD surgery rates.

## Introduction

Evidence-based medicine requires that high-quality evidence be translated into widespread clinical practice. Randomised surgical trials are increasing in frequency, yet their influence on changing surgical practice is not well characterised. In this study, we investigate the changing incidence of the most common shoulder operation (arthroscopic subacromial decompression) before, during and after the conduct of two large pragmatic randomised controlled trials[1,2] and publication of a rapid recommendation[3].

Painful shoulder is common, with a prevalence (in 2000) of 2.4% of primary care consultations in the UK[4] and 4.5 million visits to physicians in 2002 in the USA[5]. Sub-acromial pain accounts for up to 70% of all shoulder pain problems[6]. Surgical intervention by arthroscopic subacromial decompression (ASAD) is a common treatment for sub-acromial pain after failed non-operative treatment: from 2006/2007 to 2016/2017 in England its use increased by 91% from 30 to 52 procedures per 100,000, and use is higher internationally (e.g. 131 per 100,000 in Finland in 2011 [7]). This use imposes a significant economic cost, costing over £1 billion in the UK alone over a ten year period [7].

This rapid increase in ASAD surgery was accompanied by observational and interventional evidence with conflicting findings[8–11]. Two multi-centre placebo surgery controlled trials were subsequently conducted and published in 2018: the CSAW trial[1], recruiting in the UK from 2012 to 2015, and the FIMPACT trial[2], recruiting in Finland from 2005 to 2013. Both did not find evidence of clinically relevant differences in primary outcomes between ASAD and diagnostic arthroscopy (i.e. placebo surgery) for patients with subacromial pain and intact rotator cuff tendons. A 2019 Cochrane review, including eight trials, then found high certainty evidence that SAD does not provide clinically important benefits[12].

Anecdotal evidence of the international response in the provision of ASAD of different health care systems to these RCT findings seems to suggest that the incorporation of research evidence into practice is varied. In the UK and within 6 months of publication of the CSAW trial, NHS England put ASAD on a list of ‘procedures of limited value’, meaning that it should only be performed in limited circumstances[13]. In Scotland, ASAD declined consistently from 2014-2018, with the largest decrease from 2017 to 2018 coinciding with the publication of the two randomised trials[14]. However, data evaluating changes in ASAD incidence since publication of the trials in the rest of the UK are not yet available.

More broadly, international comparisons of ASAD incidence rates based on recent data are not available. Data from Florida (USA), New York (USA), Western Australia and Finland indicate that ASAD is more common than in England[7], but these data are (at the most recent) from 2013. A recent study assessing the incidence of ASAD in the US found evidence of declining use from 2010 to 2018[15], but a similar analysis has not been conducted for the UK.

This study aimed to assess the impact of the existence and publication of major ASAD trials on surgical practice. The study used large routinely collected data sets from the US, Belgium and UK to examine the temporal trends in ASAD incidence and any trends in the characteristics of patients undergoing ASAD surgery.

## Methods

This study is reported according to the STROBE reporting guideline [16].

### Study design and setting

We did a retrospective study to describe ASAD incidence in Europe and the US and examined the trends in characteristics of patients undergoing ASAD. Incidence data were stratified according to age and gender, and patient characteristics were age, gender, Charlson index [17] and medicine use in the previous month (additional characteristics were examined and are given in the aggregated data on GitHub, see “Reproducibility and data availability”).

### Setting and data sources

We retrieved primary care data from the UK, US and Belgium, in October and November 2020 from patients undergoing ASAD from 2005 to 2019 for the first time in their records in a given year. Follow-up time points extended from the beginning of each calendar year to the earliest of the end of that year, death, transfer out date (i.e. the date of the patient leaving the index general practice, or changing insurance providers), and ASAD surgery.

Outpatient electronic medical records (EMR) from Belgium, UK and US were collected. We additionally collected insurance claims data for the US, because US EMR data cover a much smaller fraction of the population. US and Belgium Data were from IQVIA and UK data were from Clinical Practice Research Datalink (CPRD) Gold (ISAC application #20_000245). Databases used are summarised as follows:

- IQVIA Longitudinal Patient Data Belgium (LPD Belgium) database consists of data collected from electronic medical records and longitudinal patient database in general practice settings. Data coverage spans 2 million unique patients, 688 care sites, 15 million visits, and 140 million service records. Dates of care include 2008 forward.
- IQVIA Ambulatory EMR (AEMR) consists of longitudinal, de-identified electronic health records originating from ambulatory clients in the US. Data coverage spans 76 million unique patients with 414 million visits. The data contains detailed clinical information that captures important health outcomes such as lab test results and vital signs in an outpatient care setting. Dates of care include 2006 forward.
- IQVIA Open Claims is a large pre-adjudicated claims database aggregated from clearinghouses that process commercial insurance claims. Data coverage spans over 280 million unique patient lives and represents more than 80% of American Medical Association physicians submitting medical and pharmacy claims in the United States. Care locations include both inpatient and outpatient facilities. Dates of care include 2005 forward.
- Clinical Practice Research Datalink (CPRD) consists of anonymised primary care data collected from a network of GP practices across the UK. The data encompass 50 million patients, including 16 million currently registered patients.

Study sample size was determined by availability of data in each database: we used all available data.

### Participants

Patients were eligible for inclusion in the cohort if they had at least one visit recorded in the database in a given year and cases were defined as patients undergoing ASAD for the first time in their records in a given year. The following codes were used to identify ASAD cases and were identified by two authors with surgical expertise (AJC and JLR):

- Arthroscopic decompression of subacromial joint
- Acromioplasty of shoulder
- Partial acromioplasty
- Arthroscopic acromioplasty
- Arthroscopy, shoulder, surgical; decompression of subacromial space with partial acromioplasty, with coracoacromial ligament (ie, arch) release, when performed (List separately in addition to code for primary procedure)
- Acromioplasty or acromionectomy, partial, with or without coracoacromial ligament release

### Quantitative variables and statistical methods

Incidence rates of ASAD per 10,000 person years were calculated as cases*10,000/person years. Characteristics of patients undergoing ASAD were assessed at time of surgery and descriptive statistics were calculated per year in which surgery was conducted. There was no missing data (that we were able to determine) in the included databases.

## Results

We obtained data from CPRD for the UK and, from IQVIA, for the US (two databases: IQVIA OpenClaims and IQVIA AEMR) and Belgium (IQVIA LPD Belgium). Number of patients, person years, cases, and years included per database are summarised in Table 1. Characteristics of patients by year and database at the time of ASAD, and number of cases per year are given in the supplementary material. In the US, IQVIA OpenClaims provided a much larger sample: we therefore report results only for OpenClaims.

**Table 1:**
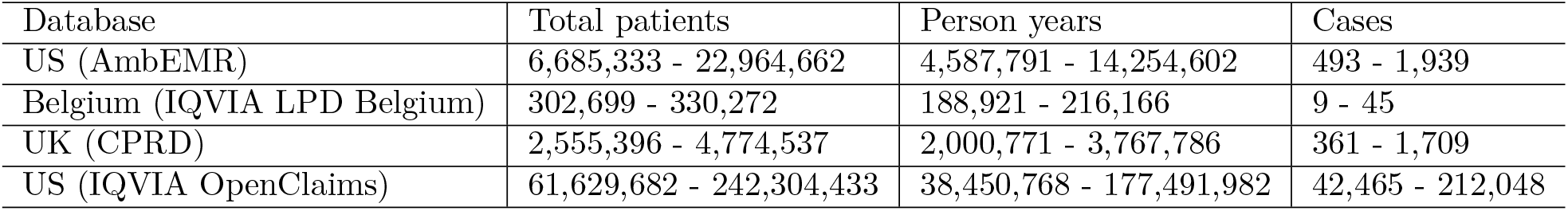
Summary of databases by total patients eligible for analysis, person years, cases, and years with at least one ASAD case. Ranges are given for each variable for the data included in this study (i.e. only for those years with at least one case)

| Database | Total patients | Person years | Cases |
| --- | --- | --- | --- |
| US (AmbEMR) | 6,685,333 - 22,964,662 | 4,587,791 - 14,254,602 | 493 - 1,939 |
| Belgium (IQVIA LPD Belgium) | 302,699 - 330,272 | 188,921 - 216,166 | 9 - 45 |
| UK (CPRD) | 2,555,396 - 4,774,537 | 2,000,771 - 3,767,786 | 361 - 1,709 |
| US (IQVIA OpenClaims) | 61,629,682 - 242,304,433 | 38,450,768 - 177,491,982 | 42,465 - 212,048 |

### Outcome data

Incidence was highly variable across databases and time (Figure 1). US incidence rates were considerably higher than those in UK and Belgium. In the UK, ASAD incidence plateaued from 2010-2014, and has consistently fallen since. There was no clear pattern in US incidence over a similar time period based on analysis of claims data. Note that historical rates in the US decrease from 2011 due to a smaller number of contributing care sites represented in the database during that period of time. The most stable data is available over the last 10 years. The number of recorded cases was extremely small in Belgium so any findings need to be interpreted with caution, but there was no clear pattern in incidence over time. Despite changes in the interim period, rates at the beginning and end of the time periods studied in the US and UK were similar (2005 - 2019).

**Figure 1:**
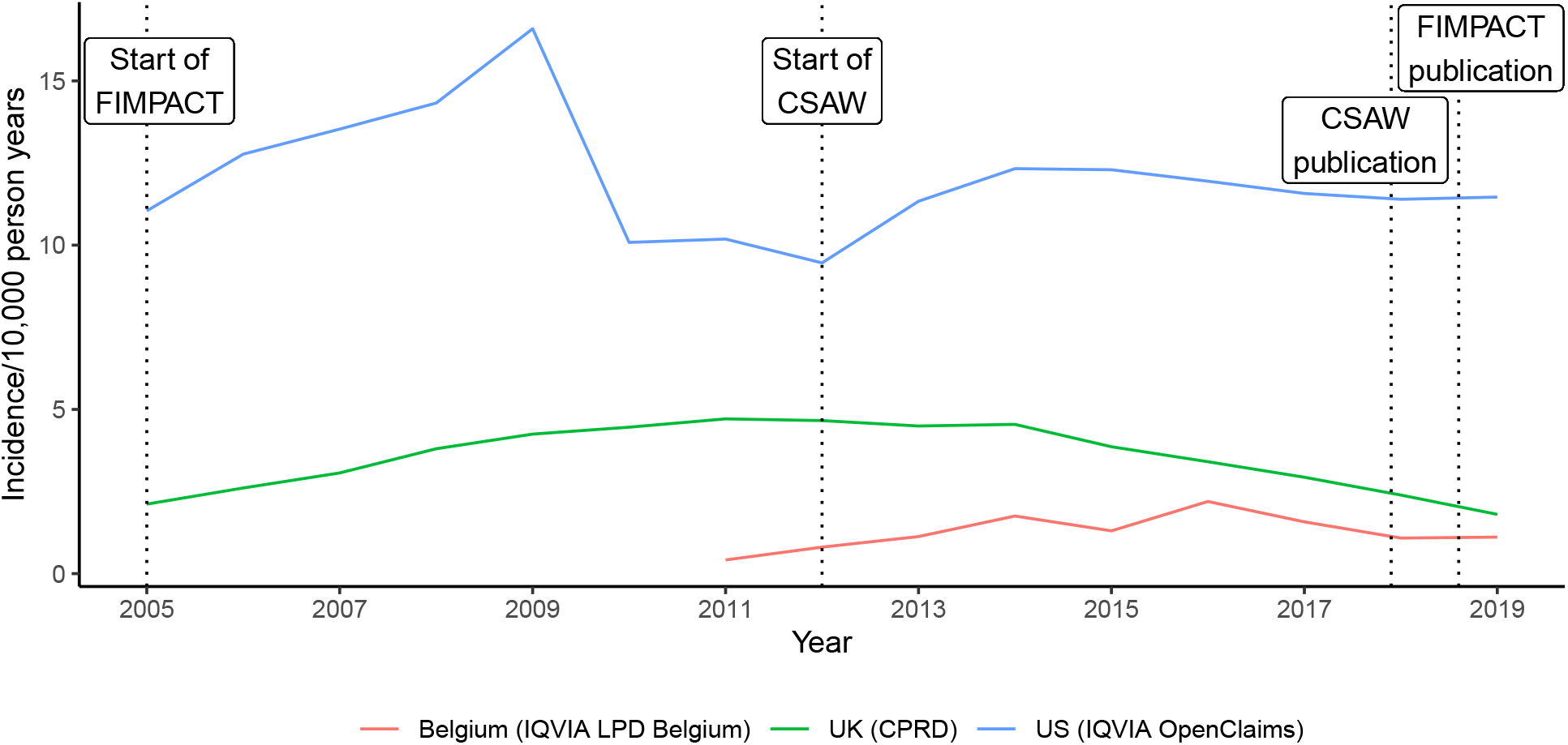
Incidence of ASAD per 10,000 person years in Belgium, UK and US. The FIMPACT[2] trial recruited in Finland from 2005 to 2013 and published in July 2018. The CSAW[1] trial recruited in the UK from 2012 to 2015 and was published in January 2018. Both trials did not find evidence of a clinically important difference compared to diagnostic arthroscopy (i.e. placebo surgery).

Changes over time were similar within different age and sex strata of the population in US and UK data (Figure 2; Belgium data are not shown because there were too few cases in several strata which could compromise anonymity). Males had higher rates of ASAD than females, the age group with the highest incidence was 50 to <65, and age <35 had the lowest incidence.

**Figure 2:**
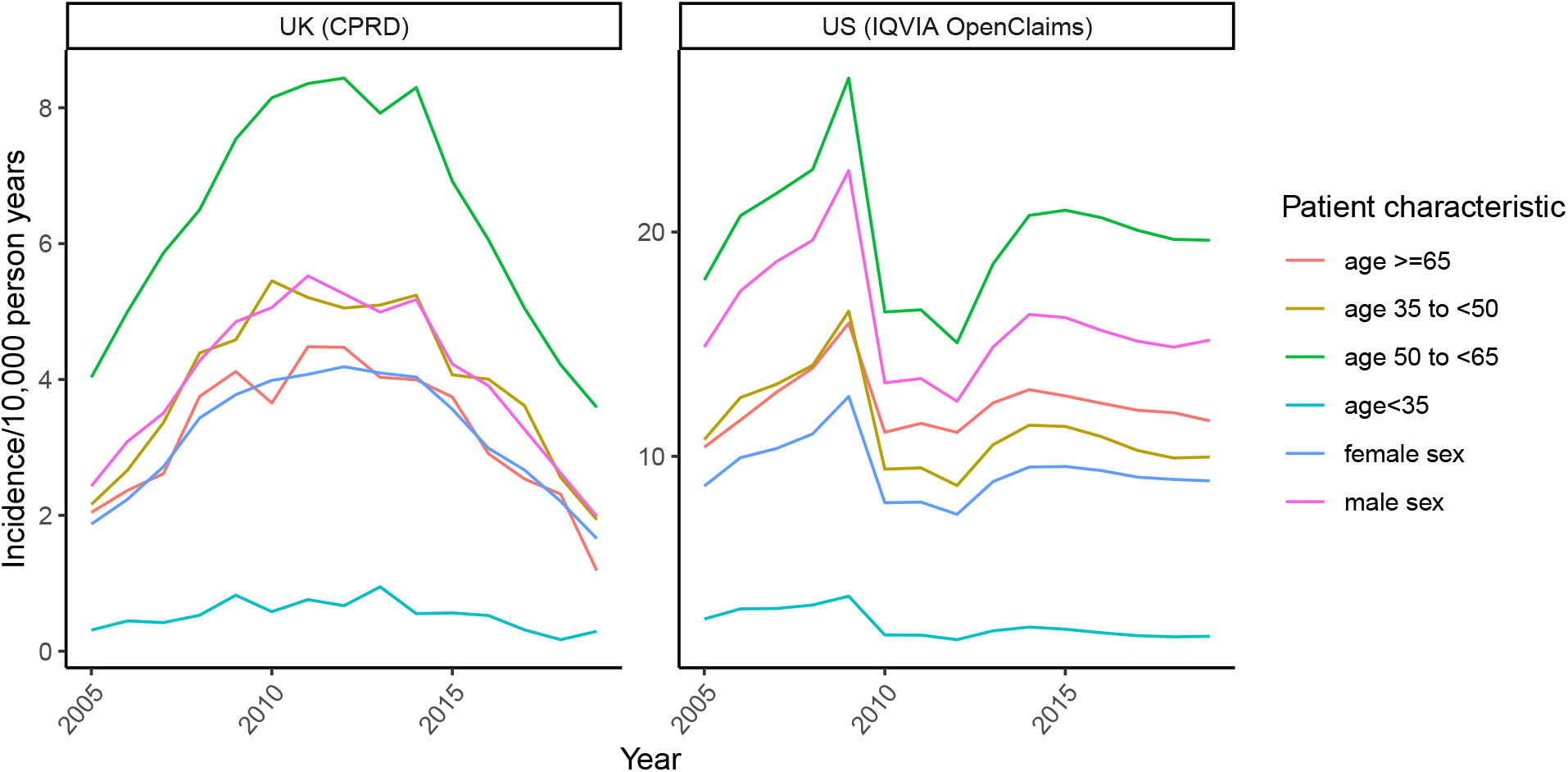
Incidence of ASAD per 10,000 person years in UK (CPRD) and US (IQVIA OpenClaims), stratified by patient characteristics. Note that the y-axis scales differ.

## Discussion

We present the most comprehensive analysis of international ASAD rates to date and the first UK-wide analysis of ASAD incidence for shoulder impingement including data from 2018 and 2019, (i.e. after publication of robust trials questioning the effectiveness of ASAD). The analysis shows that UK incidence has fallen since a peak in 2011 while there is no clear pattern for the US. US rates were higher than UK rates throughout the study period and remain high.

### Comparisons to other literature

UK data are consistent with patterns observed in previous studies, though US data contrast with a recent study. Hospital episode statistics data from 2000/2001 to 2009/2010 showed increasing rates of ASAD use in England[18]. UK incidence rates in this study increased from 2005 to 2010 by a similar magnitude. ASAD incidence in Scotland decreased from 2014 to 2018[14], a finding observed UK-wide in our study. A recent analysis of US claims data indicated that from 2010 to 2018 ASAD incidence declined by 39.6% from 118.0 to 71.3 per 100,000[15]; we find ASAD incidence rates in a similar range but increasing by 12.9% from 10.1 to 11.4 per 10,000 person years over the same period. Real world data have different rates of capture, and there is potential subjectivity in case definitions that may contribute to variability in the the rates reported.

### Interpretation

UK ASAD rates show a clear and consistent downward trend while US rates remain high (11.5/10,000 person years in US in 2019 vs. 1.8/10,000 person years in UK) despite lack of evidence indicating, for certain shoulder pathology groups, effectiveness over placebo surgery. The decline in ASAD incidence in UK from 2011 to 2019 coincides with the conduct and publication of CSAW (conducted 2012 to 2015, published 2018)[1], FIMPACT (conducted 2005 - 2013, published 2018) [2] and a Cochrane review (published 2019) of combined study data for ASAD[12]. Interestingly, a large proportion of the decrease in ASAD happened before any results of the trials were available, potentially suggesting that the awareness over the uncertainty in the effectiveness of ASAD raised by running of the national trial contributed to the reduction, in what could be termed an “ongoing RCT effect”. A paper highlighting the rapid spread of ASAD for shoulder impingement in the absence of evidence of benefit may also have contributed to the decline[18], though cannot explain it fully as it was published in 2014. It seems most plausible that a combination of these factors is responsible.

Reasons for the lack of change in rates in the US are unclear. It is possible that any “ongoing RCT effect” is specific to countries or populations in which trials are conducted, and that results of surgical RCTs take more time to change practice internationally than nationally, despite the international dissemination of results. However, data from Sweden suggest a decrease in ASAD similar to that observed in our study, and the results of the trials are rapidly informing clinical guidelines (personal communication, Stefan Lohmander). Clinicians in countries such as the US may not view the results of the studies conducted in UK and Finland to be generalisable to their practice; however, the patient cohorts recruited to these trials are clearly defined in the trial publications and are not unique to the country of origin.

### Limitations

Database coverage and reporting of procedures changes over time, so some variation in ASAD incidence may not be attributable to real changes in ASAD surgery rates. It is challenging to address these sources of error; however, we have included data from some of the largest and most representative sources available and these limitations are applicable to any research using them. In the US data, there was discordance between incidence rates in the two data sources, with rates lower from IQVIA AEMR. Lower rates most likely reflect limited capture of these procedures in an outpatient setting. EMR data have been shown in other contexts to substantially underestimate care and utilisation[19]. We therefore included IQVIA Open Claims data only in our results.

Few cases were recorded in Belgium, which could reflect low incidence or could reflect that ASAD was not captured in the data. The latter seems more plausible since the database, LPD Belgium, largely consists of outpatient data, which is not designed to capture surgery. Regardless, the number of cases was too low to draw meaningful conclusions about trends in patient characteristics or incidence. Also, we tried but were unable to retrieve incidence data for Germany, France, and Italy with the instantiated cohort definitions.

Certain codes used to identify the cohort may result in the inclusion of cases that are not arthroscopic (i.e. are open) SAD or not ASAD specifically for shoulder impingement. Acromioplasty or acromionectomy, partial, with or without coracoacromial ligament release, for example, could potentially represent open SAD. However, open SAD is much less common, for example in the US comprising only 5% of SAD surgeries performed[15]. ASADs performed as part of other shoulder operations like rotator cuff repair or resection of calcific deposit may not have been excluded with the used codes. There is inevitably a trade-off between sensitivity and specificity of codes; with two experienced shoulder surgeons helping in our cohort definition, we feel the codes used were reasonable.

## Conclusions

We found evidence that the impact of the conduct and publication of two large ASAD surgical RCTs differed between the UK and the US. In the UK, ASAD rates fell both during and after completion of a large national trial. This highlights the successful engagement of surgeons and healthcare commissioners in adopting the results of the trial, and supports the ongoing commitment in the UK to fund and conduct surgical RCTs to address treatment uncertainties. While such evidence should translate internationally, it has not, and further work is needed to understand the barriers or concerns around the international uptake of high quality evidence.

## Data Availability

This article was written in RMarkdown and can be reproduced from the publicly available aggregate data and code provided on GitHub (https://github.com/worcjamessmith/ASAD_rates_public). Some data could not be shared because it contained cells with low counts which could compromise anonymity.

## Acknowledgements

The research was supported by the National Institute for Health Research (NIHR) Oxford Biomedical Research Centre (BRC). DPA is funded through a NIHR Senior Research Fellowship (Grant number SRF-2018-11-ST2-004). The funder played no role in the study. The views expressed in this publication are those of the author(s) and not necessarily those of the NHS, the National Institute for Health Research or the department of Health.

## Reproducibility and data availability

This article was written in RMarkdown and can be reproduced from the publicly available aggregate data and code provided on GitHub (https://github.com/worcjamessmith/ASAD_rates_public). Some data could not be shared because it contained cells with low counts which could compromise anonymity. RStudio[20] and several R[21] packages[22–28] were used.

