## Supplementary material for "International impact of large multi-centre surgical trials of arthroscopic subacromial decompression"

Table 1: Characterisation of patients undergoing ASAD in Belgium (IQVIA LPD Belgium), UK (CPRD) and US (IQVIA OpenClaims), data

| Year | Age (SD) | Gender (% female) | Charlson index (SD) | Medicines previous month (IQR) | Cases |
|---|---|---|---|---|---|
| **Belgium (IQVIA LPD Belgium)** | | | | | |
| 2011 | 58.44 (17.60) | 44.44 | 1.11 (2.38) | 6 (3-7) | 9 |
| 2012 | 61.30 (15.05) | 40.00 | 0.70 (1.16) | 4 (2-6) | 17 |
| 2013 | 52.77 (12.54) | 61.54 | 0.77 (1.02) | 3 (2-7) | 24 |
| 2014 | 57.70 (15.45) | 50.00 | 1.17 (1.94) | 3 (1-6) | 36 |
| 2015 | 59.23 (12.69) | 67.50 | 0.88 (1.42) | 4 (1-5) | 27 |
| 2016 | 55.95 (12.63) | 59.65 | 0.74 (1.44) | 3 (1-5) | 45 |
| 2017 | 59.60 (13.75) | 53.49 | 0.70 (0.77) | 4 (1-7) | 32 |
| 2018 | 60.46 (12.98) | 61.54 | 0.62 (0.44) | 5 (2-8) | 22 |
| 2019 | 60.68 (14.36) | 64.00 | 1.16 (2.46) | 3 (1-6) | 21 |
| **UK (CPRD)** | | | | | |
| 2005 | 55.41 (11.94) | 49.82 | 0.53 (1.12) | 2 (0-5) | 761 |
| 2006 | 55.02 (12.34) | 47.86 | 0.51 (0.94) | 2 (0-5) | 948 |
| 2007 | 54.83 (11.98) | 49.06 | 0.65 (1.08) | 2 (0-5) | 1155 |
| 2008 | 55.33 (12.49) | 50.50 | 0.72 (1.24) | 2 (0-5) | 1413 |
| 2009 | 55.12 (12.50) | 49.15 | 0.68 (1.12) | 2 (0-6) | 1596 |
| 2010 | 54.80 (12.05) | 49.72 | 0.72 (1.23) | 2 (0-5) | 1651 |
| 2011 | 55.55 (12.03) | 48.70 | 0.74 (1.21) | 3 (0-6) | 1709 |
| 2012 | 55.73 (11.91) | 50.00 | 0.73 (1.14) | 3 (0-6) | 1687 |
| 2013 | 55.15 (12.18) | 50.92 | 0.76 (1.20) | 3 (0-6) | 1581 |
| 2014 | 55.46 (11.52) | 49.12 | 0.75 (1.12) | 3 (1-6) | 1485 |
| 2015 | 55.72 (12.02) | 50.28 | 0.75 (1.26) | 3 (0-6) | 1150 |
| 2016 | 55.04 (11.61) | 47.89 | 0.80 (1.27) | 2 (0-5) | 871 |
| 2017 | 55.07 (11.44) | 49.86 | 0.71 (1.18) | 3 (1-5) | 649 |
| 2018 | 56.33 (11.04) | 50.37 | 0.81 (1.25) | 3 (1-6) | 501 |
| 2019 | 55.39 (11.04) | 49.61 | 0.76 (1.21) | 3 (1-6) | 361 |
| **US (IQVIA OpenClaims)** | | | | | |
| 2005 | 54.83 (11.96) | 48.19 | 0.57 (1.25) | 0 (0-0) | 42465 |
| 2006 | 55.06 (12.08) | 47.78 | 0.62 (1.27) | 0 (0-0) | 49418 |
| 2007 | 55.39 (12.25) | 46.74 | 0.67 (1.32) | 0 (0-0) | 58170 |
| 2008 | 55.74 (12.41) | 46.73 | 0.73 (1.40) | 0 (0-0) | 71444 |
| 2009 | 55.58 (12.42) | 46.35 | 0.82 (1.43) | 0 (0-0) | 114409 |
| 2010 | 55.80 (12.45) | 45.99 | 0.94 (1.55) | 3 (0-6) | 139231 |
| 2011 | 56.07 (12.53) | 45.51 | 1.02 (1.63) | 4 (1-7) | 143463 |
| 2012 | 56.41 (12.65) | 45.52 | 1.11 (1.67) | 4 (2-7) | 141712 |
| 2013 | 56.14 (12.58) | 45.11 | 1.15 (1.73) | 5 (3-9) | 182021 |
| 2014 | 56.12 (12.47) | 44.53 | 1.23 (1.82) | 6 (3-11) | 204645 |
| 2015 | 56.22 (12.42) | 44.68 | 1.36 (1.90) | 7 (3-11) | 209402 |
| 2016 | 56.48 (12.34) | 45.20 | 1.49 (2.03) | 7 (3-12) | 212048 |
| 2017 | 56.83 (12.21) | 45.27 | 1.60 (2.11) | 7 (3-12) | 204810 |
| 2018 | 57.21 (12.22) | 45.58 | 1.71 (2.18) | 7 (3-12) | 199974 |
| 2019 | 57.50 (12.17) | 45.25 | 1.77 (2.24) | 7 (3-12) | 187183 |
